# mRNA vaccine boosting enhances antibody responses against SARS-CoV-2 Omicron variant in patients with antibody deficiency syndromes

**DOI:** 10.1101/2022.01.26.22269848

**Authors:** Ofer Zimmerman, Alexa Michelle Altman Doss, Paulina Kaplonek, Laura A. VanBlargan, Chieh-Yu Liang, Rita E. Chen, Jennifer Marie Monroy, H. James Wedner, Anthony Kulczycki, Tarisa L. Mantia, Caitlin C. O’Shaughnessy, Hannah G. Davis-Adams, Harry L. Bertera, Lucas J. Adams, Saravanan Raju, Fang R. Zhao, Christopher J. Rigell, Tiffany Biason Dy, Andrew L. Kau, Zhen Ren, Jackson Turner, Jane A. O’Halloran, Rachel M. Presti, Daved H Fremont, Peggy L. Kendall, Ali H. Ellebedy, Galit Alter, Michael S. Diamond

## Abstract

Patients with primary antibody deficiency syndromes (PAD) have poor humoral immune responses requiring immunoglobulin replacement therapy. We followed PAD patients after SARS-CoV-2 vaccination by evaluating their immunoglobulin replacement products and serum for anti-spike binding, FcγR binding, and neutralizing activities. Immunoglobulin replacement products had low anti-spike and receptor binding domain (RBD) titers and neutralizing activity. In COVID-19-naive PAD patients, anti-spike and RBD titers increased after mRNA vaccination but decreased to pre-immunization levels by 90 days. Patients vaccinated after SARS-CoV-2 infection developed higher responses comparable to healthy donors. Most vaccinated PAD patients had serum neutralizing antibody titers above an estimated correlate of protection against ancestral SARS-CoV-2 and Delta virus but not against Omicron virus, although this was improved by boosting. Thus, currently used immunoglobulin replacement products likely have limited protective activity, and immunization and boosting of PAD patients with mRNA vaccines should confer at least short-term immunity against SARS-CoV-2 variants, including Omicron.

## INTRODUCTION

Common variable immune deficiency (CVID) and other primary antibody deficiency syndromes (PAD) are associated with low immunoglobulin levels and impaired antibody responses to pathogens and vaccines^1,2^. Patients with these immune disorders suffer from severe and recurrent infections, autoimmunity, and are at increased risk for malignancies^3^. CVID has a prevalence of 1 to 25,000^4-6^ and is the most common primary immunodeficiency in patient registries with more than 20% suffering from this condition. CVID is not a single disease but rather a collection of hypogammaglobulinemia syndromes resulting from multiple genetic defects^7-11^. Most patients with PAD require intravenous or subcutaneous immunoglobulin replacement therapy that decreases their risk for infection^12-14^. There are more than 15 commercially available immunoglobulin products in the United States. The production of immunoglobulin replacement products takes up to one year from sample donation to distribution^15,16^. Each vial contains immunoglobulins pooled from thousands of donors^16,17^, and each manufacturer has its own plasma donors. In patients with PAD, immunoglobulin replacement is dosed every 1 to 4 weeks, depending on the route of administration.

SARS-CoV-2 is the causative agent of the global COVID-19 pandemic. From November 2019 until now, the virus has caused at least 5.5 million deaths. In the United States, the emergency use authorization has been granted for two COVID-19 vaccines (mRNA-1273, Moderna and Ad26.COV2.S, Johnson & Johnson/Janssen) and full approval has been given to one mRNA vaccine (BNT162b2, Pfizer-BioNTech). Presently, there is limited data regarding the effectiveness of mRNA or adenoviral vector vaccination against COVID-19 in CVID or PAD patients. Several studies showed variable seroconversion rates with detection of anti-spike, S1 or RBD antibodies, in 20 to 90% of PAD patients following vaccination with BNT162b2, mRNA-1273, or ChAdOx1 (Oxford-AstraZeneca)^18-21^, with better responses in those with prior history of SARS-CoV-2 infection^20,22^. Available data are limited to the initial vaccine response with no information on durability or the effect of boosting in PAD patients. Furthermore, no data has been published on the ability of PAD patient serum to neutralize authentic SARS-CoV-2 and variants, including the currently dominant B.1.1.529 (Omicron) virus. Finally, no study has reported the anti-spike, anti-RBD, or neutralization activity of immunoglobulin replacement products that PAD patients had received, to rule out the possibility that their anti-SARS-CoV-2 antibodies originated from passive immunoglobulin therapy. To address these gaps, we evaluated the effect of mRNA vaccination and boosting on serum antibody responses in PAD patients against historical SARS-CoV-2 strains and key circulating variants.

## RESULTS

### Antibody binding to spike and RBD

To begin to determine the baseline immunity afforded by antibody replacement therapy for our patient cohort, we tested 48 distinct lots of 6 different immunoglobulin products (**Table S1**) for binding to historical (Wuhan-1) spike and RBD proteins and compared these results to serum from 12 healthy donors (HD) before or 14 and 90 days post-completion of 2 doses of BNT162b2 mRNA vaccine (**Fig 1A-B**). Only one immunoglobulin product, Gamunex-C, showed anti-spike antibody titers higher than unvaccinated HD, and these values were substantially lower than those from HD 14 and 90 days post-immunization (*P* < 0.001) (**Fig 1A**). In all tested immunoglobulin products, anti-RBD titers were not different from those of unvaccinated HD (**Fig 1B**). Thus, antibody replacement products in clinical use at the time of this study (May 2021-January 2022) had remarkably low levels of anti-SARS-CoV-2 antibody and likely were derived from donations obtained before or soon after the onset of the pandemic.

**Figure 1.**
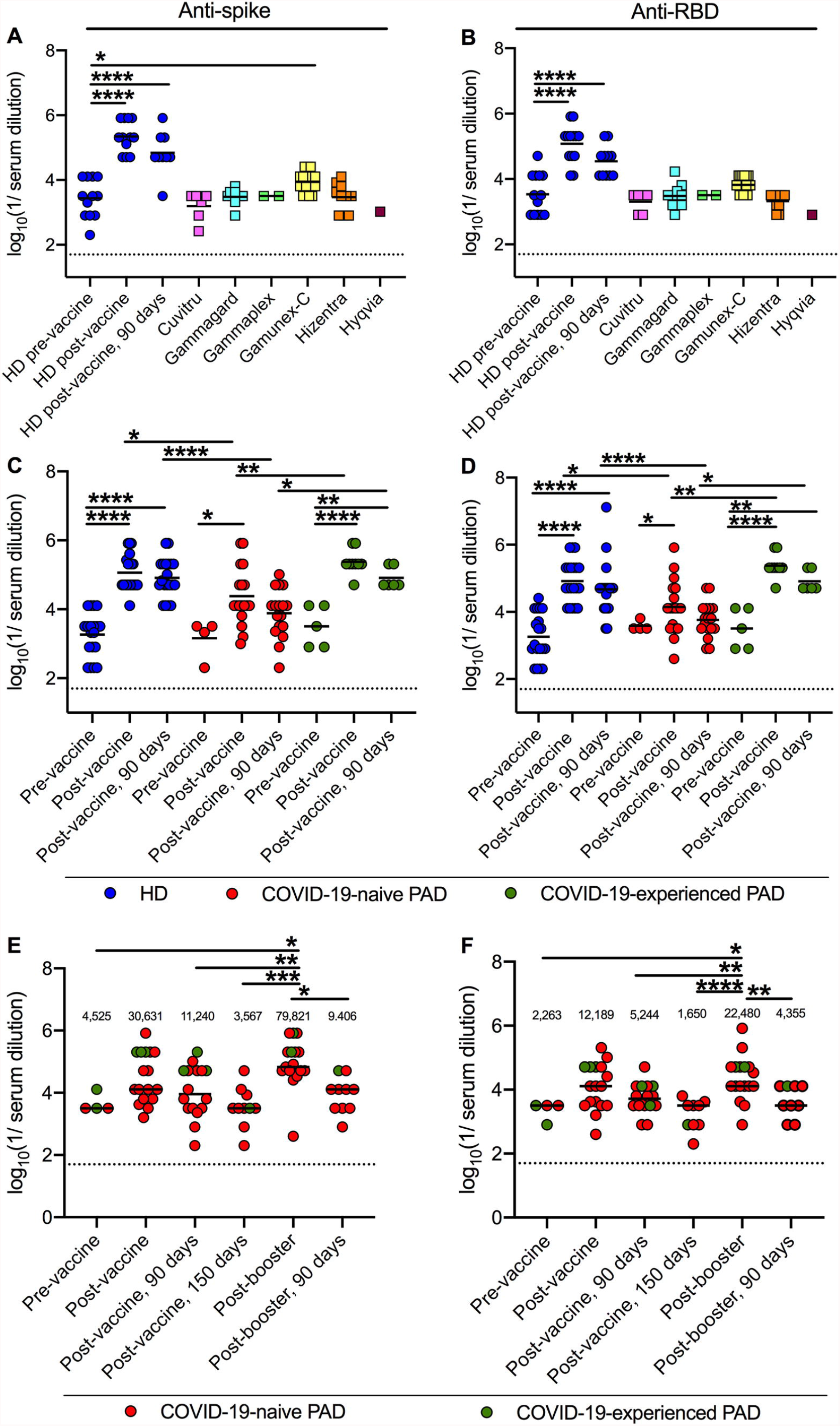
Anti-spike and anti-RBD titers following primary vaccination and boosting in PAD patients. Anti-Wuhan-1 Spike (**A**) and RBD (**B**) endpoint titers in 48 lots of 6 different immunoglobulin replacement products (squares) compared to 12 HD (blue circles) before, 14 days and 90 days post-completion of BNT162b2 vaccine series. Anti-Wuhan-1 spike (**C**) and RBD (**D**) endpoint titers in HD (n = 20; blue circles), COVID-19-naive (n = 18; red circles) and COVID-19-experienced (n = 9; green circles) PAD patients before, or 14 and 90 days post-completion of mRNA (BNT162b2, n = 19 or mRNA-1273, n = 8) vaccination series. Anti-Wuhan-1 Spike (**E**) and RBD (**F**) endpoint titers in COVID-19-naive (n = 14; red circles) and COVID-19-experienced (n = 3; green circles) PAD patients before (n = 4), 14 or 28 (n = 17), 90 (n = 16) and 150 (n = 10) days post-completion of primary mRNA (BNT162b2 n = 13, mRNA-1273 n = 2) or Ad26.COV2.S (n = 2) vaccine series, and 14 (n = 17) days and 90 (n = 10) days post-booster with mRNA vaccine (BNT162b2 n = 15; or mRNA-1273 n = 2). Dotted black line represents the limit of detection. Numbers above graphed data (**E-F**) represent the geometric mean titer (GMT) for each time point. One-way ANOVA with Dunnett’s post-test; Bars indicate mean values; Only significant differences are shown: *, *P* < 0.05; **, *P* < 0.01; ***, *P* < 0.001; ****, *P* < 0.0001).

We next compared anti-spike and anti-RBD titers of 27 PAD patients (**Table 1 and S2-S3**) who completed immunization with mRNA vaccines (BNT162b2, n = 19, mRNA-1273, n = 8) to those of 20 HD immunized with BNT162b2. Nineteen patients had CVID, with the others having hypogammaglobulinemia (n = 4) or specific antibody deficiency (n = 4) diagnoses. Nine of the 27 PAD patients had a confirmed history of prior COVID-19 infection by RT-PCR and were convalescent or recovered (COVID-19-experienced, range of 36-276 days from infection to vaccination; median 106 days; mean 130 days). Because of the study design, we obtained pre-vaccination serum samples from only a subset (9 of 30) of individuals in the PAD cohort. Notwithstanding this limitation, our analysis of immunoglobulin replacement samples (**Fig 1A-B**) suggests that individuals without a prior history of SARS-CoV-2 infection should have low, if any, levels of pre-existing anti-spike or RBD antibody. Fourteen days post-immunization, both COVID-19-naive and -experienced PAD patients as well as HD had anti-spike titers that were higher than the pre-vaccination anti-spike titers (59-fold, COVID-19-naive PAD; 53-fold, COVID-19-experienced PAD; and 55-fold, HD) (**Fig 1C**). However, at 90 days post-vaccination, PAD patients without a history of SARS-CoV-2 infection showed waning anti-spike titers to levels that were not higher than pre-vaccination baselines (**Fig 1C**). In contrast, anti-spike titers from COVID-19-experienced PAD and HD remained higher than pre-vaccination levels (17-fold, COVID-19-experienced PAD; 41-fold, HD) (**Fig 1C**). Similar findings were observed with anti-RBD titers. Fourteen days post-immunization, all groups had higher anti-RBD titers than the pre-vaccination (19-fold, COVID-19-naive PAD; 19-fold, COVID-19-experienced PAD; and 39-fold, HD) (**Fig 1D**). At 90 days post-vaccination, anti-RBD titers of COVID-19-naive PAD patients were not higher than baseline (**Fig 1D**), whereas titers of COVID-19-experienced PAD and HD had decreased but remained higher than before immunization (4-fold, COVID-19-experienced PAD; 17-fold, HD) (**Fig 1D**). Thus, vaccinated HD and COVID-19-experienced PAD patients had higher anti-spike and anti-RBD responses than vaccinated PAD patients lacking a history of SARS-CoV-2 infection (**Fig 1C-D**).

**Table 1.**
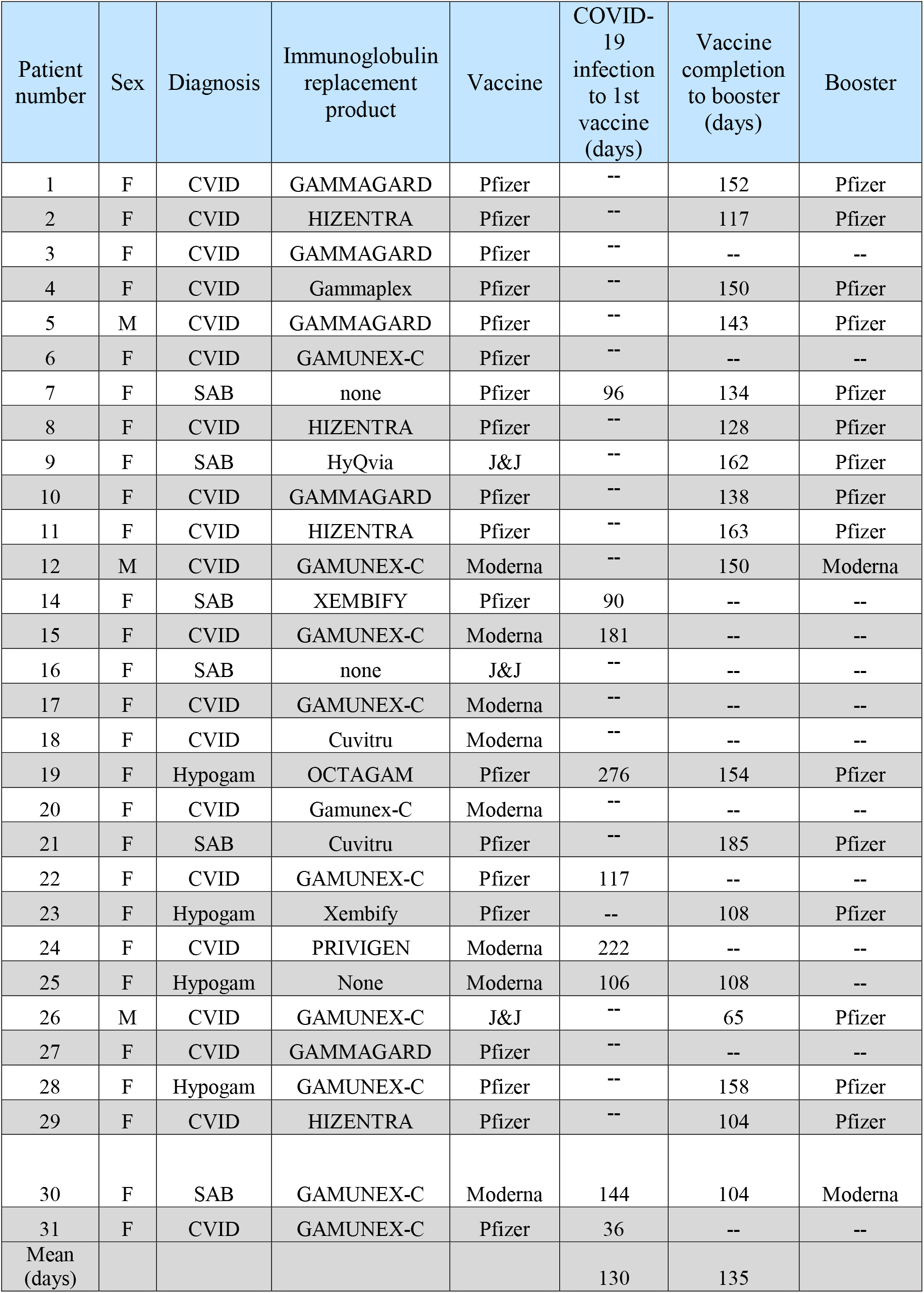
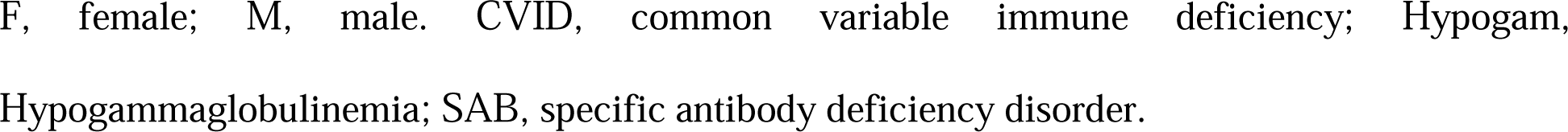
Patients characteristics.

| Patient number | Sex | Diagnosis | Immunoglobulin replacement product | Vaccine | COVID-19 infection to 1st vaccine (days) | Vaccine completion to booster (days) | Booster |
| --- | --- | --- | --- | --- | --- | --- | --- |
| 1 | F | CVID | GAMMAGARD | Pfizer | -- | 152 | Pfizer |
| 2 | F | CVID | HIZENTRA | Pfizer | -- | 117 | Pfizer |
| 3 | F | CVID | GAMMAGARD | Pfizer | -- | -- | -- |
| 4 | F | CVID | Gammaplex | Pfizer | -- | 150 | Pfizer |
| 5 | M | CVID | GAMMAGARD | Pfizer | -- | 143 | Pfizer |
| 6 | F | CVID | GAMUNEX-C | Pfizer | -- | -- | -- |
| 7 | F | SAB | none | Pfizer | 96 | 134 | Pfizer |
| 8 | F | CVID | HIZENTRA | Pfizer | -- | 128 | Pfizer |
| 9 | F | SAB | HyQvia | J&J | -- | 162 | Pfizer |
| 10 | F | CVID | GAMMAGARD | Pfizer | -- | 138 | Pfizer |
| 11 | F | CVID | HIZENTRA | Pfizer | -- | 163 | Pfizer |
| 12 | M | CVID | GAMUNEX-C | Moderna | -- | 150 | Moderna |
| 14 | F | SAB | XEMBIFY | Pfizer | 90 | -- | -- |
| 15 | F | CVID | GAMUNEX-C | Moderna | 181 | -- | -- |
| 16 | F | SAB | none | J&J | -- | -- | -- |
| 17 | F | CVID | GAMUNEX-C | Moderna | -- | -- | -- |
| 18 | F | CVID | Cuvitru | Moderna | -- | -- | -- |
| 19 | F | Hypogam | OCTAGAM | Pfizer | 276 | 154 | Pfizer |
| 20 | F | CVID | Gamunex-C | Moderna | -- | -- | -- |
| 21 | F | SAB | Cuvitru | Pfizer | -- | 185 | Pfizer |
| 22 | F | CVID | GAMUNEX-C | Pfizer | 117 | -- | -- |
| 23 | F | Hypogam | Xembify | Pfizer | -- | 108 | Pfizer |
| 24 | F | CVID | PRIVIGEN | Moderna | 222 | -- | -- |
| 25 | F | Hypogam | None | Moderna | 106 | 108 | -- |
| 26 | M | CVID | GAMUNEX-C | J&J | -- | 65 | Pfizer |
| 27 | F | CVID | GAMMAGARD | Pfizer | -- | -- | -- |
| 28 | F | Hypogam | GAMUNEX-C | Pfizer | -- | 158 | Pfizer |
| 29 | F | CVID | HIZENTRA | Pfizer | -- | 104 | Pfizer |
| 30 | F | SAB | GAMUNEX-C | Moderna | 144 | 104 | Moderna |
| 31 | F | CVID | GAMUNEX-C | Pfizer | 36 | -- | -- |
| Mean (days) |  |  |  |  | 130 | 135 |  |

F, female; M, male. CVID, common variable immune deficiency; Hypogam, Hypogammaglobulinemia; SAB, specific antibody deficiency disorder.

Seventeen PAD patients received a booster dose with an mRNA vaccine (range of 65-185 days from initial vaccine completion to booster; median 141 days; mean 135 days) (**Table 1**), and samples were obtained one to four weeks later (n = 16, range 7-27 days, median 17 days, mean 17 days; one patient had a post-booster sample drawn at day 35). Serum anti-spike and RBD titers were higher after boosting than those obtained pre-booster (5 to 9-fold compared to day 90, and 19 to 32-fold compared to day 150; **Fig 1E-F**). Although the anti-spike and RBD titers after boosting trended higher than those obtained at 14 days post primary immunization, these differences did not reach statistical significance (*P* > 0.3). Moreover, 90 days after boosting, serum titers against spike and RBD from PAD patients had decreased to levels comparable to before boosting (**Fig 1E-F**).

### Immunoglobulin subclass and Fc-gamma receptor binding

Recent studies have suggested that immunoglobulin subclass and antibody interactions with Fcγ receptors (FcγR) can contribute to protective immunity against SARS-CoV-2^23,24^. Accordingly, we evaluated serum from immunized PAD patients for their immunoglobulin subclasses (IgG1, IgG2, IgG3 and IgG4, IgA, and IgM) that bind spike proteins and domains (S, S1, S2, and RBD) from historical (Wuhan-1) and spike proteins from B.1.351 (Beta) and B.1.617.2 (Delta) SARS-CoV-2 strains (**Fig 2A-F; Fig S1**). Fourteen days after primary series immunization, COVID-19-naive PAD had lower IgG2, IgG3, and IgM levels against spike and RBD proteins of all 3 tested virus strains than vaccinated HD (**Fig 2B-C and F**). COVID-19-naive PAD patients also had lower IgG1 levels against Wuhan-1 SARS-CoV-2 spike and RBD than HD (**Fig 2A**). In comparison, vaccinated COVID-19-naive PAD patients had IgA titers against spike and RBD proteins that were similar to HD (**Fig 2E**). Vaccinated COVID-19-experienced PAD patients had lower IgG3 levels against Wuhan-1 spike, S1, and RBD (**Fig 2C**), but similar levels of IgG1, IgG2, IgA, and IgM against Wuhan-1 spike, S1, S2, and RBD and variant spike proteins compared to immunized HD (**Fig 2A-B and E-F**). Vaccinated, COVID-19-experienced PAD patients had higher levels of IgG2, IgA, and IgM against Wuhan-1 spike and RBD than vaccinated, COVID-19-naive PAD patients. COVID-19-experienced PAD patients had higher IgG3, IgA and IgM titers against S2 protein than COVID-19-naive PAD patients (**Fig 2C, E-F**). The levels of IgG4 anti-spike or RBD protein in all groups were near the limit of detection (**Fig 2D**). Although prior infection with COVID-19 in PAD patients was associated with a better vaccine response, COVID-19-naive and -experienced PAD patients had lower IgG3 responses than HD (**Fig 2C and S1**). This result suggests that class switching to IgG3 is impaired in PAD patients following infection or vaccination.

**Figure 2.**
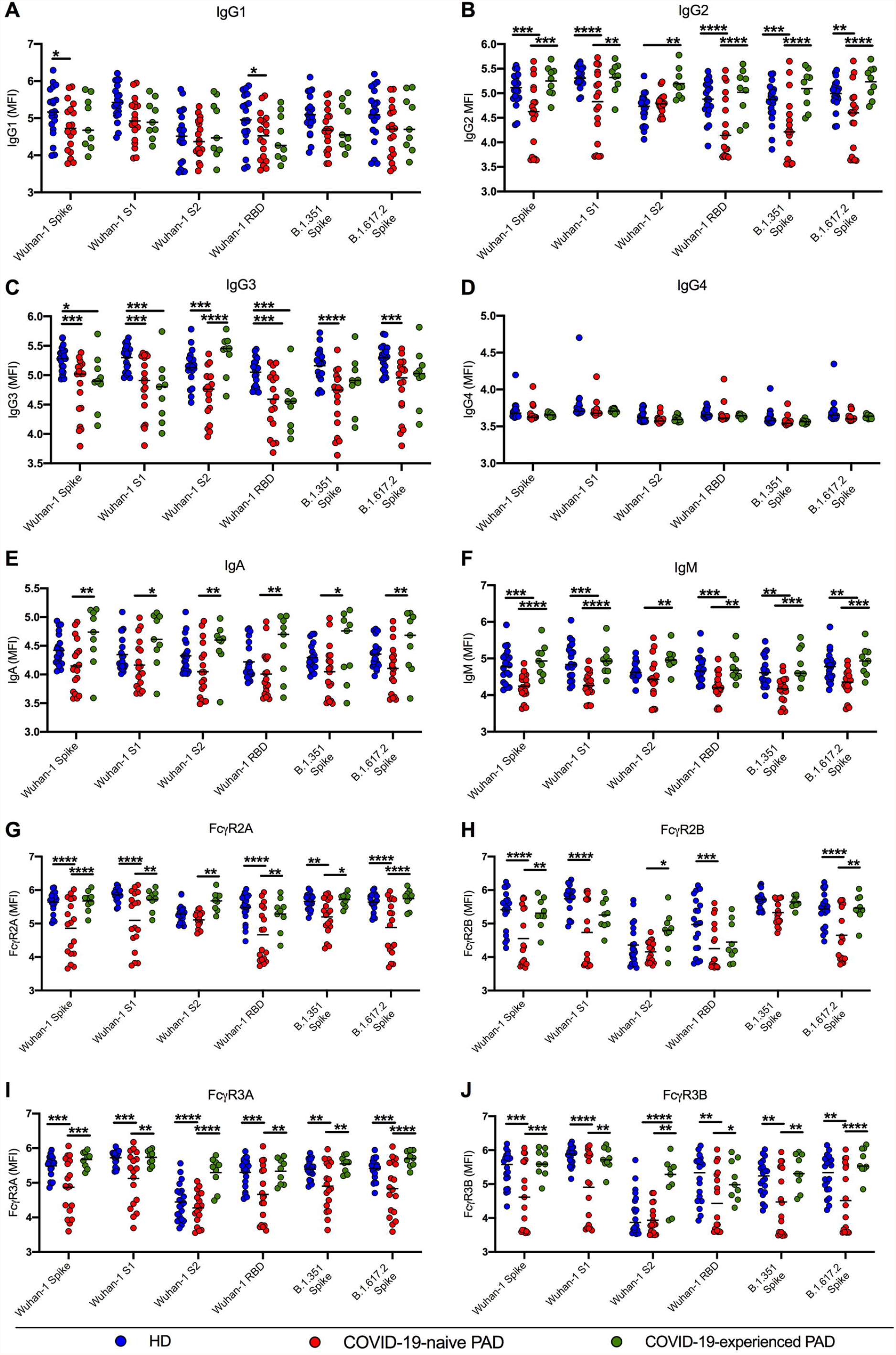
IgG subclasses and FcγR binding activity of anti-spike antibodies in serum from vaccinated PAD patients. Levels of IgG1(**A**), IgG2 (**B**), IgG3 (**C**), IgG4 (**D**), IgA (**E**), IgM (**F**), FcγR2A-binding (**G**), FcγR2B-binding (**H**), FcγR3A-binding (**I**), and FcγR3B-binding (**J**) anti-Wuhan-1 (S, S1, S2, and RBD), anti-B.1.351, and anti-B.1.617.2 spike antibodies in HD (n = 20, blue circles), COVID-19-naive (n = 18, red circles) and COVID-19-experienced (n = 9, green circles) PAD patients 14 days following the completion of mRNA vaccine series. Two-way ANOVA with Tukey post-test; mean; Only significant differences are shown: *, *P* < 0.05; **, *P*<0.01; ***, *P*< 0.001; ****, *P* < 0.0001).

Given these results, we next evaluated anti-spike and anti-RBD antibody binding to FcγRs (FcγR2A, FcγR2B, FcγR3A and FcγR3B) using a systems serology platform^25^. Vaccinated, COVID-19-naive PAD patients had lower levels of FcγR binding to anti-spike and RBD antibody than vaccinated, COVID-19-experienced PAD patients or HD (**Fig 2G-J**). FcγR2A, 3A and 3B binding was higher in serum from vaccinated, COVID-19-experienced PAD patients than vaccinated, COVID-19-naive PAD patients for all viral antigens tested (**Fig 2G, I-J and S1**). COVID-19-experienced PAD patient serum binding to FcγR2B was higher than COVID-19-naive PAD patients for Wuhan-1 and B.1.617.2 spike proteins (**Fig 2H**). FcγR2A, FcγR2B, FcγR3A and FcγR3B binding was higher in HD than COVID-19-naive PAD patients for most spike proteins (**Fig 2G-J and S1**). Serum anti-S2 responses from COVID-19-experienced PAD patients were higher than COVID-19 PAD patients for all tested FcγRs (**Fig 2G-J**). The higher levels of FcγR binding by anti-spike and anti-RBD antibodies in COVID-19-experienced compared to COVID-19-naive PAD patients after vaccination suggest that PAD patients do not have an inherent defect in producing antibodies that mediate Fc effector functions.

### Serum neutralizing antibody responses

We evaluated the functional activity of antibody preparations by performing focus reduction neutralization tests (FRNTs) with authentic SARS-CoV-2 strains and variants (WA1/2020, B.1.617.2, and B.1.1.529). We first tested neutralizing activity of the commercial immunoglobulin products (n = 17), serum from COVID-19-naive PAD patients after vaccination (n = 18), and serum from COVID-19-experienced patients after vaccination (n = 9) (**Fig 3A-B**). Fourteen of 17 different lots of immunoglobulin product tested had no appreciable neutralizing activity against WA1/2020 or B.1.617.2 at 500 μg/ml (**Fig 3A-B and S2**), which is approximately a 1/50 dilution of the mean IgG concentration measured in our PAD patients on IgG replacement therapy (9.7 mg/ml) (**Table S1 and S2**). We tested this concentration (rather than neat sample) since it corresponds to the serum dilution that is the presumed cutoff for vaccine-mediated protection^26^. Two lots of Gammunex-C and one of Hizentra had limited neutralizing activity against WA1/2020 strain at a 1/50 dilution, and only one lot (Hizentra) had inhibitory activity against B.1.617.2 at this dilution (**Fig 3A-B; S1-S2, Table S1**).

**Figure 3.**
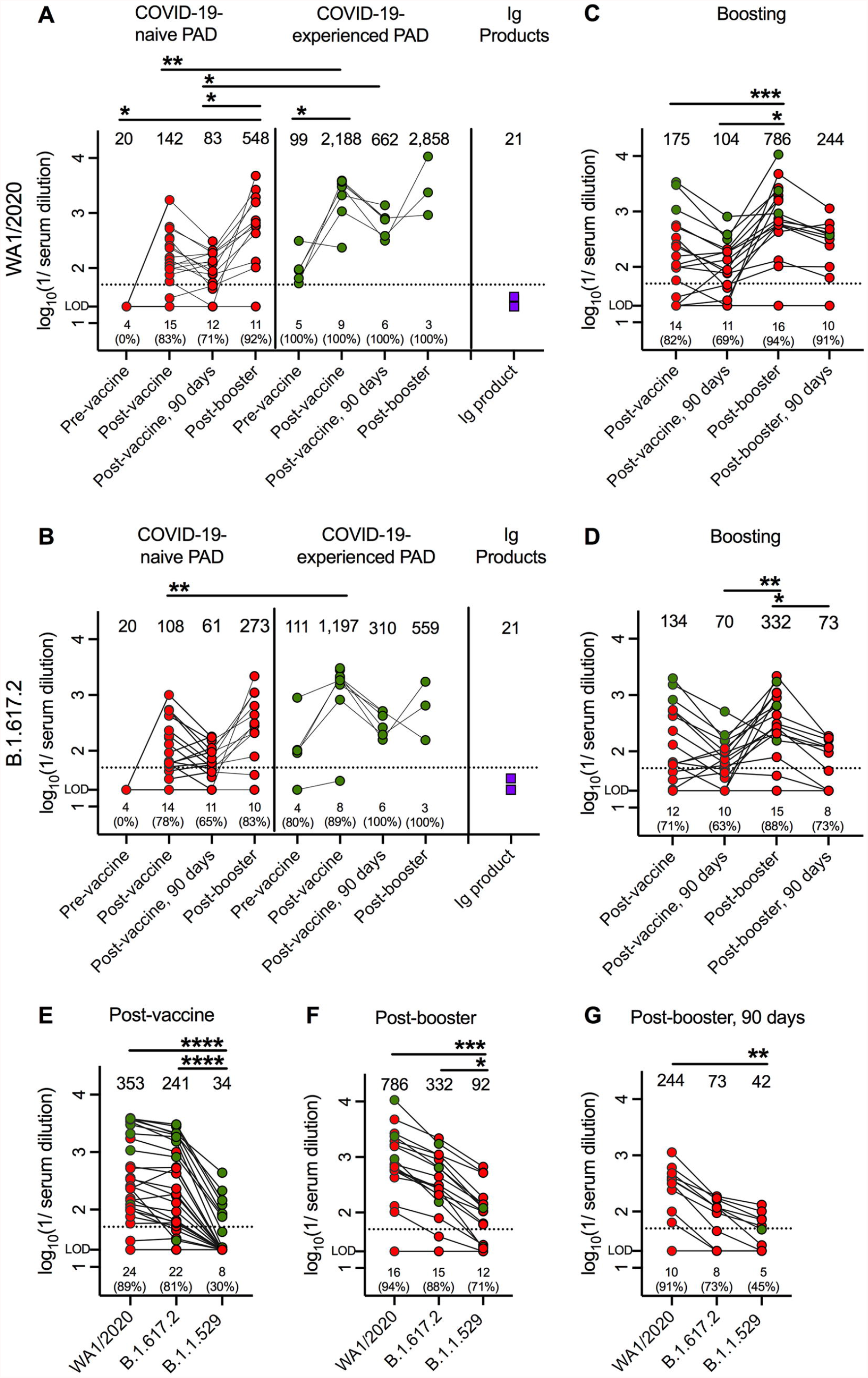
Neutralizing antibody responses in PAD patients following vaccination and bosting. Serum neutralizing activity against WA1/2020 (**A**) or B.1.617.2 (Delta) (**B**) in COVID-19-naive (red circles) and COVID-19-experienced (green circles) PAD patients before (n = 4 COVID-19-naive; n = 5, COVID-19-experienced), and 14 (n = 18, COVID-19-naive; n = 9, COVID-19-experienced) or 90 (n = 17, COVID-19-naive; n = 6, COVID-19-experienced) days following mRNA vaccination and 14 days post mRNA booster (n = 12, COVID-19-naive; n = 3, COVID-19-experienced). Neutralizing activity of immunoglobulin (Ig) replacement products (n = 17, purple squares). Effect of boosting on serum neutralization of WA1/2020 (**C**) and B.1.617.2 (**D**) in COVID-naive (n = 14, red circles) and COVID-19-experienced (n = 3, green circles) PAD patients following completion of primary mRNA (14 or 90 days post-vaccination; BNT162b2, n = 13; mRNA-1273, n = 2) or Ad26.COV2.S (28 or 90 days post-vaccination; n = 2) vaccine series and 14 or 90 days post-boosting with mRNA vaccine (BNT162b2, n = 15; or mRNA-1273, n = 2). Effect of variant strains on serum neutralizing activity of PAD patients 14 days post completion of mRNA vaccination (**E**) (n = 27 total; n = 18, COVID-19-naive, red circles; n = 9, COVID-19-experienced, green circles), 14 days post-boosting (**F**) (n = 17 total; n = 14, COVID-19-naive, red circles; n = 3, COVID-19-experienced, green circles) and 90 days post-boosting (**G**) (n = 11 total; n = 10 COVID-19-naive, red circles; n = 1, COVID-19-experienced). LOD indicates limit of detection. Dotted black line represents the presumptive protective titer as described^26^. Numbers immediately above the x axis indicate the number and percentage of patients with GMTs above 50 at each time point. Numbers above graphed data represent the geometric mean titer (GMT) for each time point. Kruskal-Wallis with Dunn’s post-test; Only significant differences are shown: *, *P* < 0.05; **, *P*<0.01; ***, *P*< 0.001; ****, *P* < 0.0001).

As expected, serum of COVID-19-naive PAD patients had no neutralizing activity against WA1/2020 prior to vaccination, whereas COVID-19-experienced patients did (**Fig 3A**). Fourteen days after immunization, 15 of 18 (83%) COVID-19-naive PAD patients had levels of serum neutralizing antibodies against WA1/2020 that are considered protective^26^ (titer > 50). By 90 days, neutralizing activity had waned, with 12 of 17 (71%) PAD patients in this group still having titers above the presumed protective threshold. Following boosting, neutralizing titers increased in 11 of 12 (92%) COVID-19-naive PAD to levels greater than 50 against WA1/2020 (**Fig 3A**). In the cohort of COVID-19-experienced PAD patients, serum neutralizing activity against WA1/2020 exceeded the protective cutoff at all tested time points in all subjects (**Fig 3A**). Indeed, 14 and 90 days post-primary immunization series, COVID-19-experienced PAD patients had serum neutralizing titers against WA1/2020 that were 20 and 8-fold higher than COVID-19-naive PAD patients, respectively (**Fig 3A**).

We repeated FRNTs with PAD patient serum and the B.1.617.2 Delta strain, which can evade neutralizing antibodies due to amino acid substitutions in the RBD^27^. Although pre-immunization serum of COVID-19-naive patients lacked inhibitory activity against B.1.617.2, serum from 4 of 5 (80%) COVID-19-experienced PAD patients neutralized B.1.617.2 before vaccination (**Fig 3B**). Fourteen and 90 days post-vaccination 14 of 18 (78%) and 11 of 17 (65%) COVID-19-naive patients, respectively had serum neutralizing titers against B.1.617.2 that were above 50. Following boosting, 10 of 12 (83%) COVID-19-naive PAD patients had neutralizing titers above 50 (**Fig 3B**). In comparison, 8 of 9 (88%), 6 of 6 (100%), and 3 of 3 (100%) of COVID-19-experienced PAD patients had serum neutralizing titers against B.1.617.2 above the presumed protective threshold at 14 and 90 days post-vaccination and 14 days post-boosting, respectively. At 14 days post-vaccination, COVID-19-experienced PAD patients had 10-fold higher serum neutralizing titers against B.1.671.2 than COVID-naive vaccinated patients (**Fig 3B**).

We next analyzed PAD patients (n = 17) who received an mRNA vaccine booster (**Fig 3C**). We included in this analysis two patients who initially received an Ad26.COV2.S vaccine (**Table 1 and Fig S3**). At 14 days and 90 post-primary vaccination, 14 of 17 (82%) and 11 of 16 (69%) patients had serum neutralization titers above 50 against WA1/2020. At 14 days post-boosting, 16 of 17 (94%) patients had neutralizing titers against WA1/2020 that exceeded 50 (**Fig 3C**), and the highest titers (GMT: 786) showed a 7-fold increase over levels at 90 days post-primary series (**Fig 3C**). Ninety days post-boosting, 10 of 11 (91%) patients had neutralization titers above 50 (**Fig 3C**). Similar findings were observed against B.1.617.2, with 12 of 17 (71%), 10 of 16 (63%), 15 of 17 (88%) and 8 of 11 (73%) PAD patients having neutralizing titers greater than 50, at 14 and 90 days post-primary immunization series or at 14 and 90 days post-boosting, respectively (**Fig 3D**). Similar to WA1/2020, the highest neutralization titers against B.1.617.2 were detected 14 days after boosting (**Fig 3D**).

The B.1.1.529 Omicron variant has >30 substitutions, deletions, and insertions in its spike protein, which jeopardizes the efficacy of vaccines designed against historical SARS-CoV-2 strains^28-30^. Accordingly, we evaluated serum neutralizing activity against B.1.1.529 in our patient cohort (**Fig 3E-F**). Fourteen days after completing the primary vaccination series, only 8 of 27 (30%) PAD patients had serum levels of neutralizing antibody above 50 against B.1.1.529 (**Fig 3E**), and only 1 of 8 patients in this group was COVID-19-naive. Fourteen days following boosting, 12 of 17 (71%) PAD patients had neutralizing titers against B.1.1.529 that exceeded 50 (**Fig 3F**). Ninety days post-boosting, 5 of 11 (45%) PAD patients had serum neutralization titers against B.1.1.529 that exceeded 50 (**Fig 3G**). The mean neutralization titer against B.1.1.529 was lower than against WA1/2020 and B.1.617.2 at 14 days after primary immunization (∼10-fold, *P* < 0.0001), 14 days post-boosting (∼4 to 8-fold, *P* < 0.05), and 90 days post-boosting (∼6-fold, *P* < 0.05), which is consistent with recent studies in immunized healthy cohorts^28-30^.

## DISCUSSION

Our findings highlight the importance of immunizing and boosting PAD patients with SARS-CoV-2 mRNA vaccines. Even though these patients have defects in their humoral responses, vaccination is an important prevention option for PAD patients against COVID-19, since immunoglobulin replacement products in use at the time of this study had limited inhibitory activity, and several of the commercially available monoclonal antibody therapeutics lose substantial neutralizing capacity against variants including B.1.1.529^31-33^. Although two doses of mRNA vaccines induce serum neutralizing antibodies in most PAD patients that presumably would protect against historical and B.1.617.2 variant, PAD patients without a history of SARS-CoV-2 infection had little to no serum neutralizing activity against B.1.1.529 (Omicron) following completion of a primary vaccination series. However, an mRNA vaccine booster in PAD patients increased anti-B.1.1.529 responses in most individuals, although the serum levels of neutralizing antibodies waned over time. Our findings support recent Center for Disease Control and Prevention recommendations for a three-dose primary mRNA vaccine series that also includes a booster dose 5 months later in moderately or severely immune suppressed indivdiuals^34^.

In patients with no prior history of COVID-19 infection, the immune response to two doses of mRNA vaccine was lower in magnitude and less durable than in HD or PAD patients with a history of infection. Of note, the increase in serum neutralizing titers following boosting was higher than the increase in anti-spike and anti-RBD titers (4.5-fold compared to 2-fold). This observation highlights the utility of performing antibody neutralization assays in addition to spike or RBD binding assays for assessing the quality of humoral immune responses. Although further studies that sample and sequence B cells in blood from PAD patients are needed, we speculate that B cells from at least some PAD patients undergo antibody maturation after infection, vaccination, and boosting.

One limitation of our study is the heterogeneity in the PAD patient cohort, which included those with CVID, hypogammaglobulinemia, or specific antibody deficiency. Although this was a limitation of the cohort available for study, we did not observe substantive differences between the patient subgroups. Instead, the most significant differences in antibody response to mRNA vaccines were between patients that had or lacked a prior history of SARS-CoV-2 infection.

Many of our PAD patients who historically had poor immune responses to bacterial and other protein antigens (*e*.*g*., *Streptococcus pneumoniae* polysaccharides, tetanus toxoid, and diphtheria toxin) as part of their initial immune workup (**Table S3**) responded to mRNA vaccines. The basis for this difference remains unclear, although it could be due to the unique adjuvant properties of the lipid nanoparticle or *in vitro*-synthesized mRNA^35-37^. In comparison, although numbers in our cohort were clearly small (n = 3), we detected little to no antibody response at 35, 60 or 90 days after immunization of COVID-naive PAD patients with the Ad26.COV2.S adenoviral-vectored vaccine (**Fig S3**). Because PAD is a heterogeneous clinical entity, with many of the genetic defects unknown^7-11^, certain classes of adjuvants or antigens may overcome specific deficiencies and promote B cell responses, albeit at lower levels than healthy counterparts. Our data suggest that the mRNA platform may have utility for vaccination of PAD patients. That said, their less durable response, lower level of anti-spike and anti-RBD IgG3, and lower levels of complement-fixing and FcγR–engaging antibodies, suggest that more frequent boosters may be required to establish and maintain protective immunity.

Our study also showed that many of the immunoglobulin replacement products currently used have low levels of inhibitory anti-SARS-CoV-2 antibodies. This may be due to the long lead time required for collection from donors, purification, and testing. Neutralization assays performed by one manufacturer and by us showed low inhibitory activity against Wuhan-1 and less activity against SARS-CoV-2 variants^38^. The three products we identified with some activity against Wuhan-1 and B.1.617.2 had titers that likely would not confer protection against B.1.1.529 given the more extensive antibody evasion by this strain^31,32,39,40^. Indeed, neutralizing titers were below the presumed protective cutoff in the 4 COVID-19-naive PAD patients who donated pre-vaccination blood samples, even though all had received immunoglobulin replacement every 3 to 4 weeks before study enrollment. It is unclear when commercially available products will have sufficient levels of specific and neutralizing anti-SARS-CoV-2 antibodies to protect PAD patients. Further binding and neutralization studies are warranted once anti-SARS-CoV-2 antibodies become more widespread in plasma pools. While many PAD patients might be eligible for long-acting combination monoclonal antibody prophylaxis (*e*.*g*., Evusheld [AZD7442]) against COVID-19, recent studies showed substantial (∼33-fold) losses in potency against B.1.1.529 Omicron virus. Thus, for now, immunization of PAD patients with mRNA vaccines that includes a booster may be the most effective way to induce a protective antibody response against SARS-CoV-2 and its variants.

## Data Availability

All data reported in this paper will be shared upon request. Any additional information required to reanalyze the data reported in this paper is available from the lead contact upon request.

## Acknowledgments

This study was supported by grants and contracts from NIH: R01 AI157155, U01 AI151810, and 75N93019C00051 [all to M.S.D.], U01AI141990, U01AI150747, HHSN272201400006C, HHSN272201400008C, and 75N93019C00051 [all to A.H.E.], and R01 DK084242 [to P.L.K.]. The study also was supported by a VA Merit Award (BX002882 [to P.L.K]). This study utilized samples obtained from the Washington University School of Medicine COVID-19 biorepository, which is supported by the NIH/National Center for Advancing Translational Sciences (UL1 TR002345). The WU353 and WU368 studies were reviewed and approved by the Washington University Institutional Review Board (approval no. 202003186 and 202012081, respectively). The authors also express their gratitude to all study participants.

## Author Contributions

O.Z. designed the study, wrote the study protocol, performed experiments, analyzed the data, and supervised the project. A.L.T.D. enrolled subjects, collected demographic and clinical data, performed experiments, and analyzed data. L.A.V, C.L. and R.E.C designed and performed FNRT experiments and analyzed data. P.K and H.L.B designed and performed Luminex profiling. P.K analyzed Luminex data. J.M.M and C.J.R collected demographic and clinical data, enrolled patients, and provided patient care. H.J.W, And.K., T.B.D, Ant.K. and Z.R. provided patient care. T.L.M wrote the study protocol, managed IRB compliance, enrolled patients, and processed patient samples. C.C.O collected patient demographic and clinical data, enrolled patients, and processed samples. H.G.D performed SARS-ELISA CoV-2 spike and RBD protein expression experiments. L.J.A and D.H.F generated crucial reagents. S.R. planned experiments and analyzed data. F.Z. designed the study and analyzed data. A.H.E and J.T contributed samples from the healthy donor cohort. J.A.O and R.M.P wrote and maintained the Institutional Review Board protocol, recruited and phlebotomized participants and coordinated sample collection of healthy donors. P.L.K supervised the project. GA designed experiments and analyzed data. M.S.D planned experiments and analyzed data. O.Z. and M.S.D. wrote the initial draft, with all other authors providing editorial comments.

## SUPPLEMENTAL FIGURE LEGENDS

**Figure S1. Systems serology signature of COVID-19-naive and COVID-19-experienced PAD patients compared to HD, Related to Figure 2.**

Composite polar plots depicting the mean level of -SARS-CoV-2-specific antibody features: (A) anti-Wuhan-1 spike, S1, S2 and RBD (B) and anti-B.1.351 or B.1.617.2 spike antibodies in HD (n = 20), COVID-19-naive PAD patients (n = 18) and COVID-19-experienced PAD patients (n = 9) 14 days following the completion of mRNA vaccine series. The size of the wedge indicates the magnitude of the mean value. The colors represent specific immunoglobulin subclass or class: IgG1, IgG2, IgG3, IgA and IgM, or specific FcγR binding: FcγR2A, FcγR2B, FcγR3A or FcγR3B.

**Figure S2. Most tested immunoglobulin replacement products have low levels of neutralizing activity, Related to Figure 3**. The relative infection rate of Vero-TMPRSS2 cells following pre-incubation of 10^2^ FFU of WA1/2020 (**A**) or B.1.617.2 (**B**) with serially-diluted immunoglobulin replacement products starting at 500 μg/ml.

**Figure S3. Immune response to primary Ad26.COV2.S vaccination and a SARS-CoV-2 mRNA boosting**. Anti-Wuhan-1 spike (**A**) and RBD (**B**) endpoint titers in COVID-19-naive PAD patients (n = 3; red circles) before, or 28, 60, 90, and 150 days after one dose of Ad26.COV2.S vaccine and 14 and 90 days post mRNA booster. Serum neutralizing activity against WA1/2020 (**C**) or B.1.617.2 (Delta) (**D**) in COVID-19-naive (n = 3, red circles) patients before and 28 or 90 days following Ad26.COV2.S vaccination and 14 or 90 days after mRNA booster. LOD indicates limit of detection. Solid blue line represents the upper limit of 95% CI of unvaccinated HD anti-spike (**A**) or anti-RBD (**B**) end point titer pre-vaccination. Dotted black line (**C-D**) represents the presumptive protective titer as described^26^. Numbers above graphed data (**A-D**) represent the geometric mean titer (GMT) for each time point. Bars (**A-B**) indicate mean values.

## METHODS

### Patients and samples

The study was approved by the Institutional Review Board of Washington University School of Medicine (Approval # 202104138). Patients were identified by a medical record search for PAD, and their records were reviewed to confirm their diagnosis and verify they met the inclusion criteria. COVID-19 vaccination status was reviewed, and subjects were contacted if they were within the vaccination window or not yet immunized. Laboratory values (**Table S2-S3**) were acquired based on review of patient history and records and were not performed as part of this study. Reference values and ranges of specific tests are based on data obtained at the time of original sampling from patient records.

Inclusion criteria included males and females over 18 years of age, health care provider-documented PAD syndrome including common variable immunodeficiency (CVID), specific antibody deficiency, or hypogammaglobulinemia, and the ability to give informed consent. Entry criteria also included receipt of a SARS-CoV-2 vaccine within 14 days of enrollment, receipt of the second dose of mRNA vaccine (Moderna mRNA-1273 or Pfizer BNT162b2) within 28 days of the first visit, or receipt of one dose of adenoviral-vector vaccine (J&J Ad26.COV2.S) within 35 days of initial visit. Exclusion criteria included participation in an investigational study of SARS-CoV-2 vaccines within the past year, history of HIV infection, an active cancer diagnosis, treatment with immunosuppressive medications, history of hematologic malignancy, treatment with anti-CD20 monoclonal antibody, receipt of live-attenuated vaccine within 30 days or any inactivated vaccine within 14 days of SARS-CoV-2 vaccination, blood or blood product donation within 30 days prior to study vaccination, and planned blood donation at any time during or 30 days after the duration of subject study participation.

469 charts were reviewed, and 160 subjects were contacted. A total of 30 adults (27 females, 3 males) with PAD met eligibility requirements and agreed to enroll in the study (see **Table 1**); we note a sex-bias in the enrollees from our PAD cohort, which is not typical for the disease itself. Ages ranged from 20 to 82, with an average age of 48.4 years old. Twenty PAD patients had CVID, six had specific antibody deficiency, and four had hypogammaglobulinemia. Twenty-seven of these subjects had received immunoglobulin replacement therapy before and during the study period from 9 different products. Nineteen subjects received the BNT162b2, 8 received mRNA-1273, and three received Ad26.COV2.S vaccines. Of the 30 subjects, 9 were diagnosed with a prior SARS-CoV-2 infection with a positive nasal swab RT-PCR test, and one received treatment with an anti-SARS-CoV-2 monoclonal antibody (bamlanivimab) 90 days prior to study enrollment.

All subjects had one mandatory post-vaccine blood sample collection with optional pre-vaccine and follow-up visits at days 60, 90, and 150 (±14 days) after vaccination. The optional pre-vaccination blood sample was collected up to 14 days before receiving vaccine. For subjects who received a two-dose series of mRNA vaccines, the first post-vaccination blood collection occurred 7 – 28 days after the second dose. For subjects receiving the Ad26.COV2.S single-dose vaccine, the first post-vaccination blood sample was collected 21-35 days after immunization. Since the study was non-interventional, patients were informed if they mounted an immune response to the vaccine, but the decision to receive a booster was made between the patient and their physician. Subjects who opted for boosting provided a blood sample up to 14 days prior to receiving the booster dose, unless the subject previously provided a sample within 2 weeks as part of the optional post-vaccine assessments. Subjects returned for an additional sample 7-28 days after receiving the booster (range 7-27 days, median 17 days, mean 17 days. One patient had her post-booster sample drawn at day 35), with a second post-booster visit and sample collection at 90 ±14 days. Immunoglobulin replacement product vials that were used in PAD patients were collected at each study visit and/or post-infusion at the Washington University Allergy and Immunology Division infusion centers.

### Healthy donor controls

Immunocompetent healthy donor volunteer blood samples were obtained as previously described^41^.

### SARS-CoV-2 spike and RBD protein expression

Genes encoding SARS-CoV-2 spike protein (residues 1-1213, GenBank: MN908947.3) and RBD (residues 319-514) were cloned into a pCAGGS mammalian expression vector with a C-terminal hexahistidine tag. The spike protein was stabilized in a prefusion form using six proline substitutions (F817P, A892P, A899P, A942P, K986P, V987P)^42^, and expression was optimized with a disrupted S1/S2 furin cleavage site and a C-terminal foldon trimerization motif (YIPEAPRDGQAYVRKDGEWVLLSTFL)^43^. Expi293F cells were transiently transfected, and proteins were purified by cobalt-affinity chromatography (G-Biosciences) as previously described^44,45^.

### ELISA

Maxisorp ELISA (Thermo Fisher) plates were coated with SARS-CoV-2 Whuan-1 spike or RBD (2 µg/ml) overnight in sodium bicarbonate buffer, pH 9.3. Plates were washed four times with PBS and 0.05% Tween-20 and blocked with 4% BSA in PBS for 1 h at 25°C. Plates were then incubated with 50 μL of patient and healthy donor serially-diluted samples (starting at 1/50) in 2% BSA PBS, for 2 hours at 25°C on a shaker. Immunoglobulin replacement products were diluted to 10 mg/ml (average patient IgG level) and then treated as described above. Plates were washed with PBS and 0.05% Tween-20 and incubated with horseradish peroxide (HRP)-conjugated goat anti-human IgG (H + L) (1:2000 dilution, Jackson ImmunoResearch) for 1 h at room temperature. After washing, plates were developed with 100 μL of 3,3’-5,5’ tetramethylbenzidine substrate (Thermo Fisher) for 120 sec and fixed with 50 μL of 2N H_2_SO_4_. Plates were read at 450 nM using a microplate reader (Synergy H1; BioTek).

### Luminex profiling

Serum samples were analyzed by a customized Luminex assay to quantify the levels of antigen-specific antibody subclasses and FcγR binding profiles, as previously described^46,47^. Briefly, SARS-CoV-2 antigens were coupled to magnetic Luminex beads (Luminex Corp) by carbodiimide-NHS ester-coupling (Thermo Fisher). Antigen-coupled microspheres were washed and incubated with plasma samples at an appropriate sample dilution (1:100 for antibody isotyping and 1:1000 for all low-affinity FcγRs) overnight in 384-well plates (Greiner Bio-One). Unbound antibodies were washed away, and antigen-bound antibodies were detected by using a PE-coupled detection antibody for each subclass and isotype (IgG1, IgG2, IgG3, IgA1, and IgM; Southern Biotech), and FcγRs were fluorescently labeled with PE before addition to immune complexes (FcγR2a, FcγR2b, FcγR3a, FcγR2b; Duke Protein Production facility). After one hour of incubation, plates were washed, and flow cytometry was performed with an iQue (Intellicyt), and analysis was performed on IntelliCyt ForeCyt (v8.1). PE median fluorescent intensity (MFI) is reported as a readout for antigen-specific antibody titers.

### Focus reduction neutralization test

Serial dilutions of immunoglobulins products or sera were incubated with 10^2^ focus-forming units (FFU) of different strains of SARS-CoV-2 for 1 h at 37°C. Antibody-virus complexes were added to Vero-TMPRSS2 cell monolayers in 96-well plates and incubated at 37°C for 1 h. Subsequently, cells were overlaid with 1% (w/v) methylcellulose in MEM supplemented with 2% FBS. Plates were harvested 30 h later by removing overlays and fixed with 4% PFA in PBS for 20 min at room temperature. Plates were washed and sequentially incubated with an oligoclonal pool of SARS2-2, SARS2-11, SARS2-16, SARS2-31, SARS2-38, SARS2-57, and SARS2-71^48,49^ anti-spike antibodies and HRP-conjugated goat anti-mouse IgG (Sigma) in PBS supplemented with 0.1% saponin and 0.1% BSA. SARS-CoV-2-infected cell foci were visualized using TrueBlue peroxidase substrate (KPL) and quantitated on an ImmunoSpot microanalyzer (Cellular Technologies).

### Quantification and statistical analysis

Statistical significance was assigned using Prism Version 9 (GraphPad) when *P* < 0.05. Statistical analysis was determined by one-way ANOVA with Dunnett’s post-test, two-way ANOVA with Tukey post-test or a Kruskal-Wallis with Dunn’s post-test. The statistical tests, number of independent experiments, and number of experimental replicates are indicated in the Figure legends.

