## Supplementary figures and images for "mRNA vaccine boosting enhances antibody responses against SARS-CoV-2 Omicron variant in patients with antibody deficiency syndromes"

### Figure S1

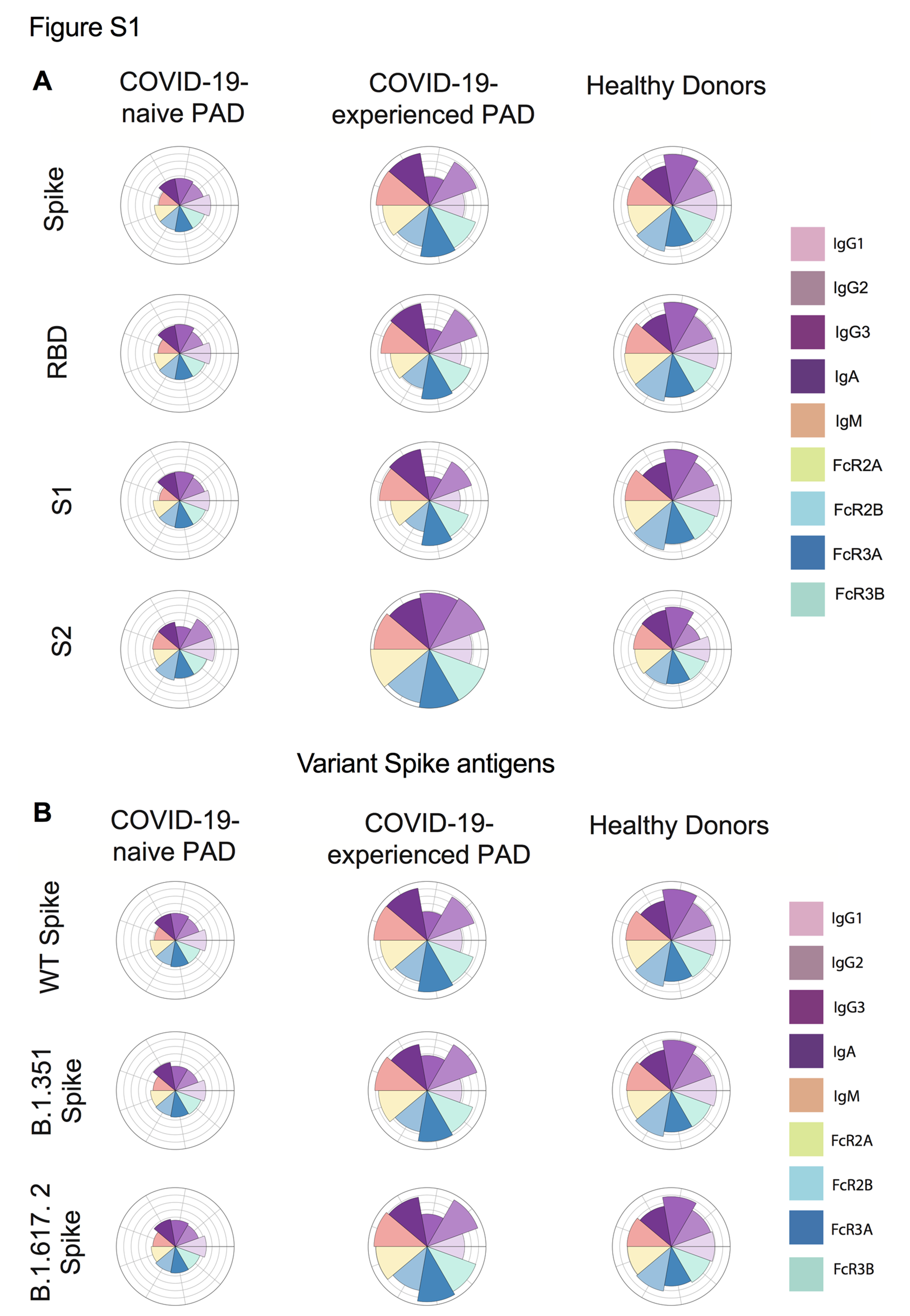

### Figure S2

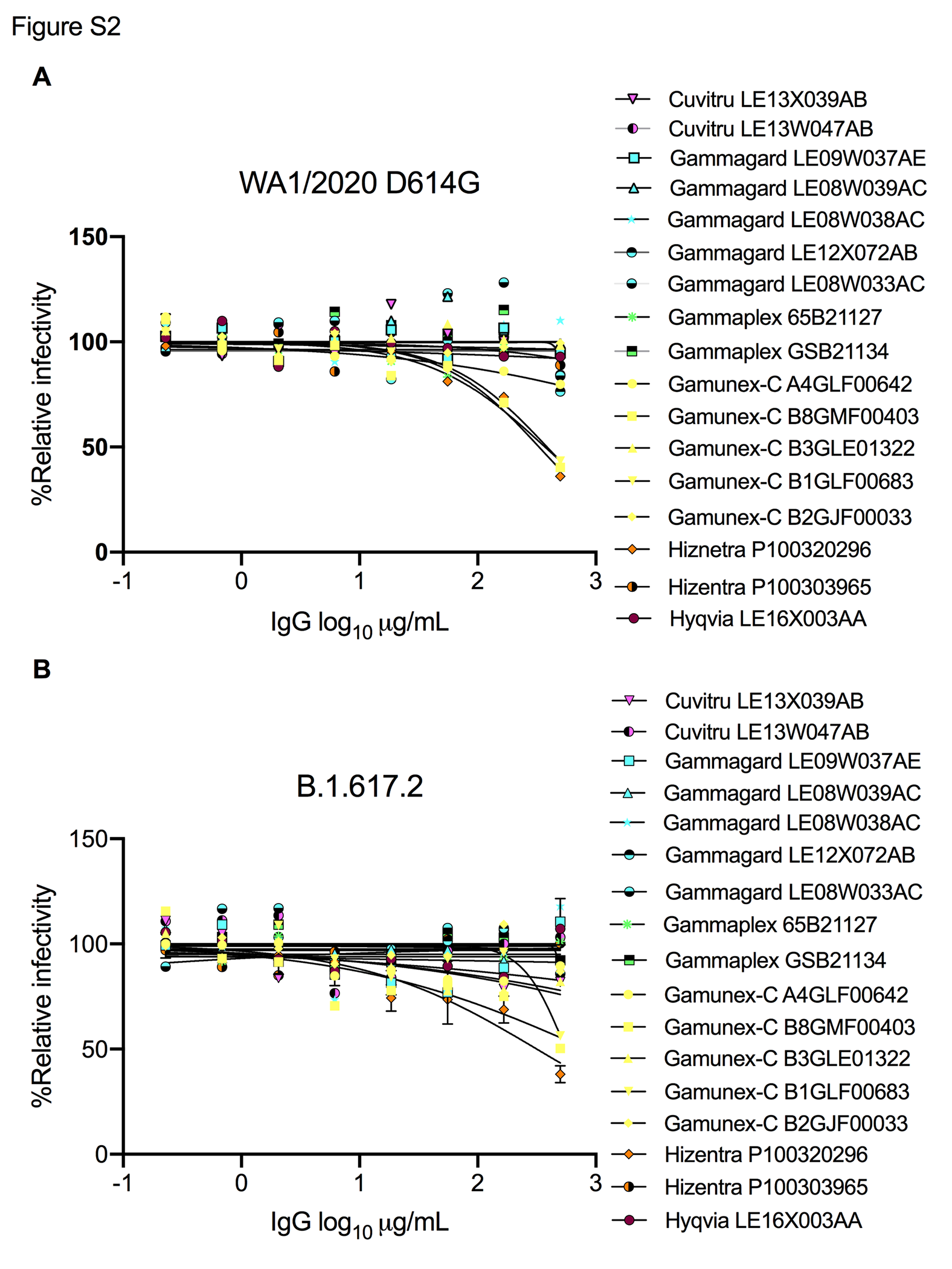

### Figure S3

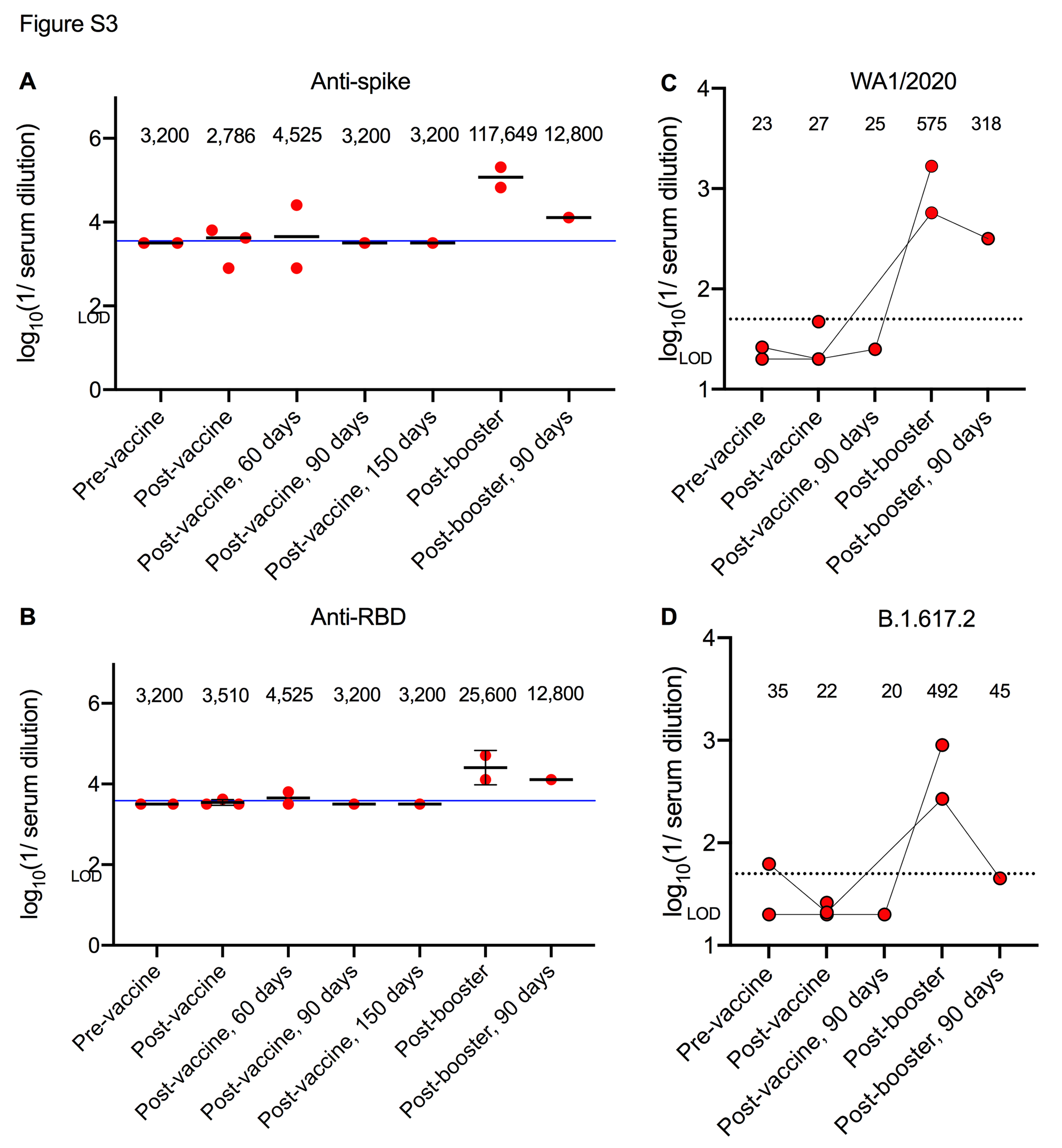
