## Supplementary material for "mRNA vaccine boosting enhances antibody responses against SARS-CoV-2 Omicron variant in patients with antibody deficiency syndromes": Table S1

**Table S1. Lot numbers of tested immunoglobulin replacement products.**

| Immunoglobulin Replacement Products | | | | | | | |
| --- | --- | --- | --- | --- | --- | --- | --- |
| Manufacturer | Lot Number | Expiration Data | Patient sample | Anti-spike End Point Titer | Anti-RBD End Point Titer | IC50 WA1/2020 D614G | IC50 B.1.617.2 |
| Gamunex-C | B2GKF00113 | -- | -- | 3200 | 3200 |  |  |
| Gamunex-C | A4GLF00642 | -- | 110 | 6400 | 6400 | 20 | 20 |
| Gamunex-C | B8GMF00363 | -- | -- | 6400 | 12800 |  |  |
| Gamunex-C | B8GLF00653 | -- | -- | 6400 | 6400 |  |  |
| Gamunex-C | A1GKF00072 | -- | -- | 12800 | 6400 |  |  |
| Gamunex-C | B8GMF00403 | 4/20/21 | 126 | 25600 | 12800 | 27 | 20 |
| Gamunex-C | B8GMF00503 | -- | -- | 5200 | 12800 |  |  |
| Gamunex-C | B3GKF00183 | -- | -- | 12800 | 6400 |  |  |
| Gamunex-C | B8GJE00743 | 10/4/23 | 128 | 6400 | 3200 |  |  |
| Gamunex-C | A4GLF00663 | -- | -- | 12800 | 6400 |  |  |
| Gamunex-C | B3GLE01322 | -- | 115 | 3200 | 3200 | 20 | 20 |
| Gamunex-C | B8GJE00733 | -- | 110 | 6400 | 12800 |  |  |
| Gamunex-C | A4GMF00383 | 4/5/24 | 119 | 12800 | 6400 |  |  |
| Gamunex-C | B1GLF00683 | 6/4/24 | 106, 110, 130 | 25600 | 6400 | 26 | 20 |
| Gamunex-C | B8GKF00243 | 4/9/24 | 126, 130 | 25600 | 12800 |  |  |
| Gamunex-C | B3GJE00633 | 9/6/23 | 119 | 3200 | 3200 |  |  |
| Gamunex-C | B2GJF00033 | 1/8/24 | 106, 130 | 6400 | 3200 | 20 | 20 |
| Gamunex-C | B86KF00233 | 4/8/24 | 130 | 12800 | 6400 |  |  |
| Gamunex-C | -- | -- | 128 | 12800 | 12800 |  |  |
| Gammaplex | 65B21127 | 4/24/24 | 104 | 3200 | 3200 | 20 | 20 |
| Gammaplex | GSB21134 | 4/27/24 | 104 | 3200 | 3200 |  |  |
| Hyqvia | LE16X003AA | 11/19/23 | 109 | 1056 | 800 | 20 | 20 |
| Gammagard | LE12X072AB | 2/22/24 | 101 | 3200 | 2112 |  |  |
| Gammagard | C21G036AAA | 3/23/24 | 101 | 3200 | 1600 |  |  |
| Gammagard | LE12X043AB | 3/17/24 | 101 | 3200 | 6400 |  |  |
| Gammagard | LE09W037AE | 11/30/22 | 105, 112 | 3200 | 16896 | 20 | 20 |
| Gammagard | LE09W016AB | -- | -- | 3200 | 1600 |  |  |
| Gammagard | LE08W039AC | 12/13/22 | 127 | 6400 | 3200 | 20 | 20 |
| Gammagard | LE08W026AD | -- | 112 | 4224 | 3200 |  |  |
| Gammagard | LE08W033AC | -- | 112 | 4224 | 4224 |  |  |
| Gammagard | LE08W010AD | 3/31/22 | 127 | 800 | 800 |  |  |
| Gammagard | LE08W038AC | 12/6/22 | 105, 112, 127, 131 | 2112 | 3200 | 20 | 20 |
| CUVITRU | LE13X039AB | -- | 118 | 2112 | 3200 | 20 | 20 |
| Manufacturer | Lot Number | Expiration Data | Patient sample | Anti-spike End Point Titer | Anti-RBD End Point Titer | IC50 WA1/2020 D614G | IC50 B.1.617.2 |
| CUVITRU | LE13X032AB | -- | 118 | 3200 | 3200 |  |  |
| CUVITRU | LE13W047AB | 5/5/23 | 121 | 264 | 800 |  |  |
| CUVITRU | LE13W097AC | 10/28/23 | 121 | 800 | 800 |  |  |
| CUVITRU | LE13W070AB | 8/29/23 | 121 | 3200 | 3200 |  |  |
| CUVITRU | LE13X012AB | 2/10/24 | 121 | 3200 | 3200 |  |  |
| Hizentra | P100303965 | 7/25/23 | 102, 111, 129 | 800 | 1600 |  |  |
| Hizentra | P100317965 | -- | 129 | 800 | 800 |  |  |
| Hizentra | P100320296 | 9/16/23 | 108, 129 | 8448 | 3200 | 30 | 32 |
| Hizentra | P100317961 | 9/1/23 | 108, 129 | 4224 | 3200 |  |  |
| Hizentra | P100301497 | -- | 129 | 6400 | 1600 |  |  |
| Hizentra | P100298818 | 7/7/23 | 102 | 3200 | 800 |  |  |
| Hizentra | P100308202 | 8/8/23 | 102 | 12800 | 3200 |  |  |
| Hizentra | P100305419 | 8/1/23 | 102 | 800 | 3200 |  |  |
| Hizentra | P100295167 | -- | 102 | 3200 | 3200 |  |  |
| Hizentra | P100325992 | 9/28/23 | 108 | 3200 | 3200 |  |  |

Immunoglobulin replacement products tested in this study and their titers of anti-Spike, anti-RBD, and SARS-CoV-2 neutralization. Immunoglobulin replacement products samples were obtained at 10 mg/ml. Level of detection for neutralization is 1/20.
