## Supplementary material for "mRNA vaccine boosting enhances antibody responses against SARS-CoV-2 Omicron variant in patients with antibody deficiency syndromes": Table S2

**Table S2. PAD patient laboratory characteristics^a^**

| Patient  number | Most Recent CBC | | Lymphocyte Subpopulation at the Time of Diagnosis | | | | | Immunoglobulin levels at the Time of  Diagnosis | | | Most Recent IgG |
| --- | --- | --- | --- | --- | --- | --- | --- | --- | --- | --- | --- |
|  | WBC | ALC | CD3 | CD4 | CD8 | CD19 | CD16/56 | IgG | IgA | IgM |  |
| Normal range | 3.8 – 9.9  (1000s per mm^3^) | 1,000-3,300 (per mm^3^) | 661 – 1963 (per mm^3^) | 490 – 1294 (per mm^3^) | 187 – 781 (per mm^3^) | 110 – 488 (per mm^3^) | 76 – 467  (per mm^3^) | 700 - 1600 mg/dL | 75 - 400 mg/dL | 40 - 230 mg/dL | mg/dL |
| 1 | 5.8 | 2400 | 2118 | 1668 | 470 | 283 | 258 | 477 | 53 | 62 | 1065 |
| 2 | 4 | 1000 | 895 | 426 | 432 | 227 | 127 | 560 | 62 | 24 | 1134 |
| 3 | 4.5 | 1674 | 1450 | 934 | 456 | 187 | 180 | 566 | 140 | 106 | 828 |
| 4 | 5.1 | 1000 | 818 | 601 | 193 | 251 | 18 | <40 | <4 | <5 | 725 |
| 5 | 12 | 1500 | 974 | 384 | 487 | 280 | 221 | <300 | <10 | <25 | 947 |
| 6 | 7.2 | 1600 | -- | -- | -- | -- | -- | 432 | 89 | 95 | 830 |
| 7 | 6.5 | 1900 | 1109 | 775 | 294 | 80 | 88 | 514 | 72 | 165 | 505 |
| 8 | 9.5 | 3500 | Reported normal in immunology clinic note | | | | | 529 | 188 | 35 | 938 |
| 9 | 6.4 | 1100 | -- | -- | -- | -- | -- | 895 | 142 | 105 | 1345 |
| 10 | 4.8 | 1700 | 1632 | 859 | 738 | 184 | 59 | 1304 | 82 | 37 | 1462 |
| 11 | 5.2 | 860 | -- | -- | -- | -- | -- | 465 | -- | -- | 836 |
| 12 | 4.3 | 1000 | 1162 | 375 | 663 | 134 | 151 | 1850 | <25 | <19 | 1303 |
| 14 | 8.3 | 1900 | 1702 | 1054 | 547 | 243 | 41 | 911 | 221 | 58 | 1291 |
| 15 | 15.6 | 2400 | -- | -- | -- | -- | -- | 340 | 12 | reported nl | 1261 |
| 16 | 8 | 2400 | 1974 | 1167 | 690 | 262 | 71 | 983 | 162 | 52 | 983 |
| 17 | 5.4 | 1600 | 2194 | 1646 | 549 | 462 | 144 | 553 | 175 | 49 | 1143 |
| 18 | 7.3 | 1700 | Reported normal in immunology clinic note | | | | | 647 | <4 | <5 | 758 |
| 19 | 5 | 2200 | 2055 | 1396 | 790 | 211 | 79 | 473 | 120 | 167 | 877 |
| 20 | 8.3 | 2600 | 1848 | 1282 | 501 | 157 | 199 | 528 | 136 | 155 | 1164 |
| 21 | 6.2 | 2400 | 859 | 716 | 119 | 95 | 227 | 622 | 133 | 137 | 989 |
| 22 | 10.6 | 2720 | 3300 | 2274 | 1026 | 580 | 446 | 606 | 204 | 30 | 743 |
| 23 | 8 | 1500 | 1055 | 835 | 205 | 315 | 158 | 665 | 162 | 45 | 887 |
| 24 | 9.1 | 1190 | Reported low NK cells only in immunology clinic note | | | | | 502 | 42 | 95 | 547 |
| 25 | 4.9 | 2000 | 1837 | 1086 | 725 | 475 | 308 | 656 | 270 | 10 | 657 |
| 26 | 4.5 | 1600 | Reported low CD19 at 39, all others normal in immunology note | | | | | <200 | 66 | <25 | 860 |
| 27 | 3.8 | 1400 | 925 | 638 | 287 | 64 | 43 | 516 | <25 | 521 | 781 |
| 28 | 7.2 | 800 | 655 | 389 | 236 | 209 | 78 | 629 | 196 | 80 | 990 |
| 29 | 5 | 1420 | -- | -- | -- | -- | -- | 261 | <10 | <25 | 1240 |
| 30 | 6.27 | 1610 | -- | -- | -- | -- | -- | 814 | 99 | 198 | 1145 |
| 31 | 10.79 | 1700 | -- | -- | -- | -- | -- | 346 | <10 | <25 | 1036 |

CBC – complete blood count; WBC – white blood count, ALC – absolute lymphocyte count; Ig – immunoglobulin; dL – deciliter; Values out of the normal range are marked in red.

Empty cells (dashed lines) indicate that the test was not performed.

^a^Data acquired by review of patient records.
