## Supplementary material for "mRNA vaccine boosting enhances antibody responses against SARS-CoV-2 Omicron variant in patients with antibody deficiency syndromes": Table S3

**Table S3. Patient responses to other vaccine antigens at the time of diagnosis^a^**

| Patient | *S. pneumoniae* Titer* | Tetanus Titer  IU/ml*** | Diphtheria Titer  IU/ml*** | Genetic testing |
| --- | --- | --- | --- | --- |
| 101 | 8/23 | N/A | -- | -- |
| 102 | 7/23 | 0.51 | -- | -- |
| 103 | 10/23 | Positive | Positive | -- |
| 104 | 0/23 | Positive | -- | 3 Unrelated VUS |
| 105 | 0/23** | Negative | -- | Unrelated VUS |
| 106 | 4/13 | 4.59 | -- | -- |
| 107 | 14/23 | 0.97 | 0.09 | -- |
| 108 | 2/23 | 0.06 | 0.01 | -- |
| 109 | 6/23 | >7 | 2.34 | -- |
| 110 | 5/23 | N/A | -- | -- |
| 111 | 9/14 | 0.98 | -- | -- |
| 112 | N/A | N/A | -- | -- |
| 114 | 12/23 | 1.56 | -- | *LRBA* heterozygous variant |
| 115 | Unprotective | -- | -- | -- |
| 116 | 14/23 | 0.36 | Positive | -- |
| 117 | 11/23 | 0.01 | -- | -- |
| 118 | -- | -- | -- | -- |
| 119 | 19/23 | 0.45 | -- | PID panel negative |
| 120 | 14/23 | >2.24 | -- | -- |
| 121 | 8/23 | 0.46 | -- | -- |
| 122 | 10/23 | 0.72 | -- | -- |
| 123 | 20/23 | >2.24 | -- | -- |
| 124 | 2/24 | -- | -- | -- |
| 125 | 7/23 | 2.20 | -- | *IFIH1* heterozygous variant |
| 126 | 3/24** | 0.24 | Negative | -- |
| 127 | 0/13 | -- | -- | -- |
| 128 | 14/23 | 0.87 | 0.07 | -- |
| 129 | Non-protective | -- | -- | -- |
| 130 | 0/23 | 2.75 | -- | -- |
| 131 | -- | 1.73 | 0.35 | -- |

*Indicates the number of tested anti-*Streptococcus pneumoniae* serotypes with a level above 1.3 μg/ml. the Immune response to pneumovax vaccine/booster was considered normal when titers were higher than 1.3 μg/ml for ≥17 tested serotypes.

**Pre-pneumovax booster values; patient started on immunoglobulin replacement prior to post-booster recheck.

***A value of ≥0.01 IU/ml is considered positive. In some records, a positive notation was indicated but no value was provided.

VUS variant of unknown significance. IU – international unit. N/A – data was obtained but not available in the medical record. Empty cells (dashed lines) indicate that the test was not performed. Values out of the normal range are marked in red.

^a^Data acquired by review of patient records.
